# Low-frequency genetic variants in GAK enhance Golgi function and protect against Parkinson’s disease

**DOI:** 10.1101/2025.08.13.25333123

**Authors:** C. Calatayud, I. Fernandez-Carasa, N. Spataro, C. Mussolino, Y. Richaud-Patin, A. Faella, R. Fernández-Santiago, M. Ezquerra, T. Courtin, S. Bandres-Ciga, A. Miguez, J.M. Canals, M. Chiritoiu, V. Malhotra, A. Garrido, M.J. Marti, E. Tolosa, E. Bosch, T. Cathomen, F.H. Gage, A. Raya, A. Consiglio

**Author notes:** Correspondence to (A.R.) or (A.C.).

## Abstract

Genome-wide association studies (GWAS) have contributed significantly to unraveling the genetic bases of complex diseases such as Parkinson’s disease (PD); yet experimental evidence for causation is often elusive. Here, we hypothesized that non-manifesting carriers of a PD-causing mutation in the *LRRK2* gene could express genetic modifiers conferring disease protection. Using a pluripotent stem cell-based model, we showed that dopaminergic neurons derived from these individuals were partially protected from the disease in vitro, and that this protective effect is genetically driven. Whole-exome sequencing identified a previously unreported low-frequency variant in cyclin G-associated kinase (GAK) that was associated with a nearly nine-year delay in age at onset among LRRK2 mutation carriers in a local cohort, although replication in additional cohorts was inconclusive. To rule out inter-cohort heterogeneity, we used CRISPR/Cas9-mediated gene editing to isolate the effect of the mutation. We found that the candidate protective variant prevented neuron loss in vitro along with an improvement of several indicators endocytic-mediated transport. Together, our findings provide mechanistic insights into PD pathogenesis and actionable genetic information for the prognosis of PD patients.

**One Sentence Summary:** Investigating genetic protection against Parkinson’s disease in non-manifesting carriers of LRRK2 mutations by CRISPR/Cas9-based genome edition.

## Introduction

Parkinson’s disease (PD) is an incurable neurodegenerative disorder that results from a complex interplay between environmental and genetic factors^1,2^. A small percentage of PD cases is attributable to the inheritance of mutations in certain genes, among which *LRRK2* 6055G>A (LRRK2 G2019S) is the most frequent ^3^. Moreover, the study of LRRK2 G2019S-associated PD (L2-PD) cases serves as an adequate proxy for the sporadic disease since their clinical course is nearly indistinguishable from that of sporadic PD patients and also share similar neuropathological changes^4^. Following this rationale, several groups including ours have studied the emergence of disease-related phenotypes in dopamine neurons (DAns) derived from induced pluripotent stem cells (iPSCs) generated from L2-PD patients. Interestingly, these models recapitulated disease hallmarks such as alpha-synuclein accumulation and DAn neurodegeneration^5–7^.

However, even when L2-PD patients share the same causal mutation, their clinical presentation, including age-at-disease-onset (AAO), is variable and some LRRK2 G2019S mutation carriers never manifest disease symptoms despite advanced age^8–10^. Genome-wide association studies (GWAS) have revealed several loci influencing AAO in LRRK2 G2019S carriers, but they often failed to replicate in additional cohorts^11,12^ and importantly, genetic association does not imply causation^13,14^. In this regard, recent developments in CRISPR/Cas9-mediated genome editing of iPSC-based disease models provide the opportunity to directly test the hypothesis of causality underlying genetic association^15–18^. Here, we used this strategy in the context of non-manifesting carriers of the LRRK2 G2019S mutation and identified a low-frequency variant in cyclin G-associated kinase (GAK) that acts as a protective factor on both the appearance of disease-related phenotypes *in vitro* and delaying AAO in PD patients.

## Results

### Modeling non-manifesting carriers of LRRK2 G2019S mutation

To increase the phenotypic (and presumably genotypic) diversity for studying the effect of genetic modulators of L2-PD, we recruited three non-manifesting carriers of the LRRK2 G2019S mutation (L2-NMC). At the time of submission, participants were in the older adult age range and were assigned study-specific codes for anonymization. Detailed medical examination performed by specialized PD experts revealed no signs or symptoms of ongoing or prodromal disease^19^ (**Table S1**). Particularly predictive of pheno-conversion in asymptomatic LRRK2 G2019S carriers are imaging biomarkers such as DAT-SPECT and substantia nigra echogenicity^20,21^, both of which were normal in SP_19, SP_20 and SP_22. Therefore, we concluded that these individuals were good candidates to be considered life-long asymptomatic or, at a minimum, late converters. iPSC lines representing these L2-NMC were generated and, upon full characterization (**Fig. S1A-F**), used in the present studies alongside previously derived iPSC lines representing healthy individuals (control) and L2-PD patients^22^ (**Fig. 1A**).

**Fig. 1.**
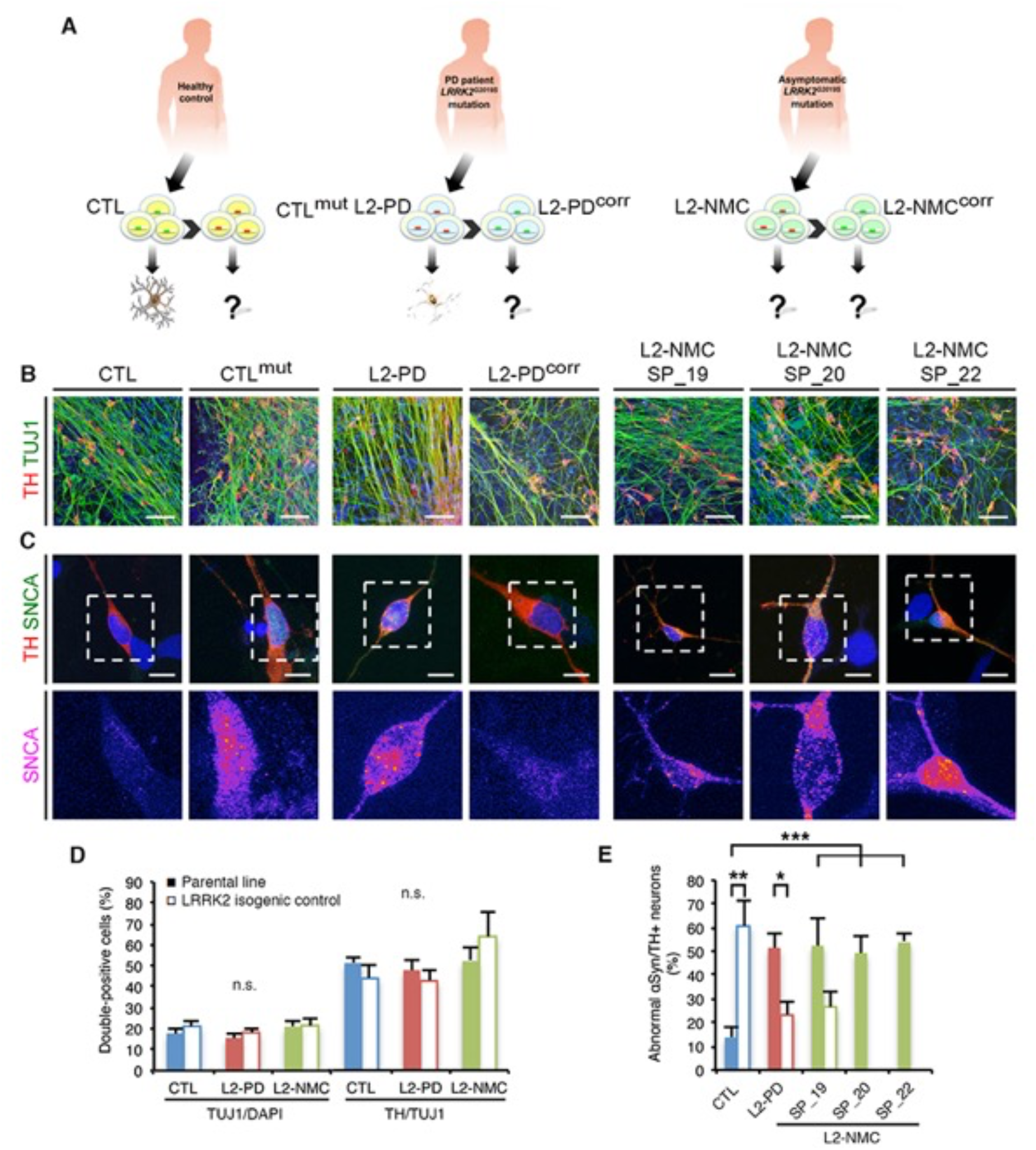
Modeling non-manifesting carriers of LRRK2 G2019S mutation. (**A**) Depiction of the overall experimental strategy; see main text for details. (**B**) Immunofluorescence staining of cell markers characteristic of DAns (tyrosine hydroxylase, TH in red) and neurons (TUJ1 in green). Nuclei were counterstained with DAPI (blue). Scale bar, 25 μm. (**C**) Immunofluorescence analyses of DAns differentiated from iPSC lines of the indicated genotypes, stained for TH (red), alpha-synuclein (SNCA, green) and DAPI (blue). The areas boxed in the upper images are shown below displaying alpha-synuclein staining only, using pseudo-coloring to facilitate visualization of staining intensity (violet tones represent low intensity whereas red to yellow tones represent high intensity). Scale bar, 15 μm. (**D**) Quantitative analysis of the percentage of neurons (cells double-stained for TUJ1 and DAPI, left set of bars) or DAns (neurons double-stained for TUJ1 and TH, right set of bars) differentiated from iPSC lines representing the indicated genotypes. Solid bars represent the parental iPSC lines and empty bars their isogenic controls. Data are averages ± s.e.m. of at least three independent experiments per genotype. Control (CTL), 2,879 DAns from SP_09; CTL^mut^, 3,178 DAns from SP_11^mut^; L2-PD, 8,248 DAns from 2 iPSC lines (SP_12 and SP_13); L2-PD^corr^, 5,461 DAns from 2 iPSC lines (SP_12^corr^ and SP_13^corr^); L2-NMC, 12,848 DAns from 3 iPSC lines (SP_19, SP_20, and SP_22); L2-NMC^corr^, 2,080 DAns from SP_19^corr^. No significant differences were found in the ability of the different iPSC lines to generate neurons [F(5.21) = 0.9115; p=0.4924] or DAns [F(5.21) = 0.5648; p=0.7259] after 21 days of differentiation. (**E**) Quantitative analysis of the percentage of DAns presenting abnormal alpha-synuclein localization at day 30. Solid bars represent the parental iPSC lines and empty bars their isogenic controls. Data are averages ± s.e.m. of two to seven independent experiments per genotype: Control, 116 DAns from 3 iPSC lines (SP_11, SP_09 and SP_17); Control^mut^, 13 DAns from SP_11^mut^; L2-PD, 61 DAns from 2 iPSC lines (SP_12 and SP_13); L2-PD^corr^, 77 DAns from 2 iPSC lines (SP_12^corr^ and SP_13^corr^); L2-NMC, 91 DAns from SP_19, 37 DAns from SP_20, and 47 DAns from SP_22; L2-NMC^corr^, 22 DAns from SP_19^corr^. Asterisks denote statistically significant differences (* p<0.05; ** p<0.01; *** p<0.001). [F(6, 22) = 7.801; p=0.0001].

We next subjected the control, L2-PD, and L2-NMC iPSC lines to a directed differentiation protocol that results in enriched populations of DAns^23^. After three weeks of differentiation, all iPSC lines generated similar numbers of DAns that co-expressed neuronal (TUJ1) and the dopaminergic marker tyrosine hydroxylase (TH) (**Fig. 1B, D**). No evident signs of neurodegeneration were observed for any of the iPSC lines at this early time-point of differentiation, as judged by the overall morphology of the neuronal network and the neurite structure of individual neurons (**Fig. 1B and Fig S2A**). Despite the absence of gross morphological anomalies, L2-PD DAns displayed abnormal accumulations of cytoplasmic alpha-synuclein compared to control DAns, in agreement with previous reports^22,24,25^, as did L2-NMC DAns (**Fig. 1C, E**). To test whether this phenotype was specific to the presence of the LRRK2 G2019S mutation, we generated isogenic iPSC controls solely differing in the correction or introduction of the mutation (**Fig. S1G-K**). Correction of the LRRK2 G2019S mutation in L2-PD or L2-NMC DAns (L2-PD^corr^ or L2-NMC^corr^, respectively) normalized the levels of alpha-synuclein to those of control DAns. Conversely, introducing the LRRK2 G2019S mutation in heterozygosis in control iPSCs (CTL^mut^) resulted in abnormal alpha-synuclein accumulation in DAns (**Fig. 1C, E**). These results indicate that the abnormal accumulation of alpha-synuclein in DAns, a PD-relevant *in vitro* phenotype previously shown to be caused by impaired proteostasis^24^, is fully penetrant with the LRRK2 G2019S mutation and, therefore, does not appear to be modified by the genetic background of control, L2-PD or L2-NMC individuals.

### Variable penetrance of neurodegeneration in L2-NMC

In addition to the abnormal accumulation of alpha-synuclein found at early culture times, L2-PD iPSC-derived DAns display increased susceptibility to neurodegeneration upon long-term culture^22,26^ or experimentally induced redox stress^25,27^. This phenotype also correlates with the presence of the LRRK2 G2019S mutation, since genetically corrected L2-PD DAns displayed percentages of apoptotic (cleaved Caspase3-positive) cells similar to controls, and control DAns engineered to carry the LRRK2 G2019S mutation underwent apoptosis at rates close to L2-PD DAns (**Fig 2 A-B**, see also^25^). Strikingly, the numbers of apoptotic DAns in L2-NMC samples were higher than those of control DAns but lower than those of L2-PD DAns cultured in parallel for 75 days (**Fig. 2A-B**). These results suggest that DAn neurodegeneration in our system, although strongly influenced by the presence of the LRRK2 G2019S mutation, appears to be modulated by the individual genetic background and exhibits incomplete penetrance in L2-NMC samples.

**Fig. 2.**
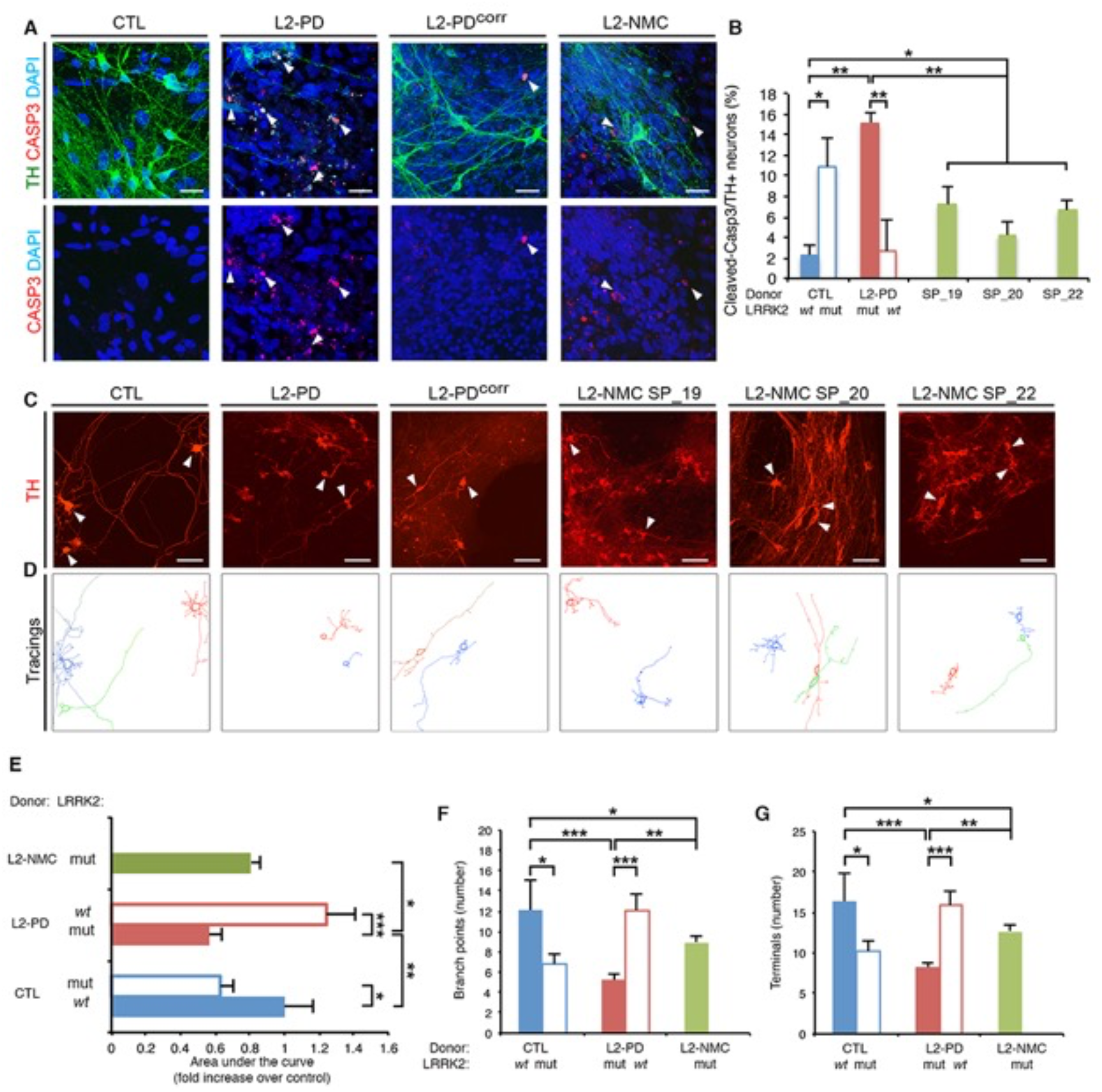
DA neurons from L2-NMC are resistant to neurodegeneration. (**A**) Immunofluorescence analyses of DAns differentiated from iPSCs of the indicated genotypes after 75 days of culture on top of mouse cortical astrocytes, stained for tyrosine hydroxylase (TH, green), the apoptosis marker cleaved Caspase-3 (CASP3, red), and DAPI (blue). Arrowheads indicate TH+/Casp3+ cells. Scale bar, 25 μm. (**B**) Quantitative analysis of the percentage of DAns staining positive for Casp3 at day 75. Data are averages ± s.e.m. of at least three independent experiments: Control (CTL), 1,064 DAns from SP_11; CTL^mut^, 320 DAns from SP_11^mut^; L2-PD, 706 DAns from 2 iPSC lines (SP_12 and SP_13); L2-PD^corr^, 1,445 DAns from 2 iPSC lines (SP_12^corr^ and SP_13^corr^); L2-NMC, 1,210 DAns from 3 iPSC lines (SP_19, SP_20 and SP_22). Asterisks denote statistically significant differences (* p<0.05; ** p<0.01). [F(6, 20) = 6.82; p=0.0005]. (**C**) Immunofluorescence analysis of DAns differentiated from iPSCs of the indicated genotypes after 75 days of culture on top of mouse cortical astrocytes, stained for TH (red) (upper panels). Arrowheads point to DAns being traced in **D**. Scale bar, 50 μm. (**D**) Neurite tracings of representative DAns for subsequent Sholl analysis. (**E-G**) Quantitative analysis of the area under the curve (AUC) of the Sholl analysis (**E**), number of branch points (**F**) and number of terminals (**G**) measured in DAns stained in **C**. Data are averages ± s.e.m. of at least three independent experiments: Control (CTL), 29 DAns from SP_11; CTL^mut^, 29 DAns from SP_11^mut^; L2-PD, 63 DAns from 2 iPSC lines (SP_12 and SP_13); L2-PD^corr^, 49 DAns from 2 iPSC lines (SP_12^corr^ and SP_13^corr^); L2-NMC, 96 DAns from 3 iPSC lines (SP_19, SP_20 and SP_22). Asterisks denote statistically significant differences (* p<0.05; ** p<0.01; *** p<0.001). ANOVA for Sholl AUC: [F(4, 261) = 6.604; p<0.0001]; for number of branches: [F(4, 261) = 6.393; p<0.0001]; for number of terminals: [F(4, 261) = 6.601; p<0.0001].

To investigate this possibility further, we quantified the shape and maintenance of the DAn neurite tree, since pathogenic *LRRK2* mutations have been associated with shortening and reduced neurite complexity in a variety of experimental models^22,25,28–30^. For this purpose, we performed Sholl analysis on DAns surviving after long-term culture and found decreased arborization complexity in L2-PD samples compared to control, as judged by the area under the curve in Sholl plots (**Fig. 2C-E and Fig. S3**). This decrease in neurite arborization complexity was dependent on the presence of the LRRK2 G2019S mutation, as isogenic controls showed the opposite trend (**Fig. 2E**). In turn, L2-NMC DAns showed, on average, neurite arborization that was of intermediate complexity, being statistically significantly more complex than L2-PD DAns but less complex than control DAns (**Fig. 2C-E**). Similarly, the numbers of terminals and branch points measured in L2-NMC DAns were significantly higher than those of L2-PD DAns, but lower than those of control DAns (**Fig. 2F-G**). It is important to note that the reduction in DAn neurite complexity detected after long-term culture (75 days) was a sign of *in vitro* neurodegeneration rather than reflecting differences in DAn differentiation or maturation, since DAn arborization analyzed at earlier time points (21 days) was consistently more complex than at 75 days of culture and showed no differences between control, L2-PD, or L2-NMC samples (**Fig. S2**). These results indicate that, under controlled environmental conditions (*in vitro* culture), DAns from L2-NMC are less susceptible to undergoing neurodegeneration than DAns from L2-PD.

### Identification of genetic modifiers in L2-NMC exomes

Our results thus far indicated that DAns from L2-NMC appeared partially protected from LRRK2 G2019S-induced neurodegeneration. Given that environmental factors were kept common across all the different cell lines *in vitro*, we reasoned that the protective mechanisms should be intrinsic to the cells being studied, either genetic or epigenetic. Since a defining feature of iPSC-based disease models is that epigenetic marks acquired by somatic cells throughout the donor’s life are virtually erased during the reprogramming process^31^, we focused on protective factors of a genetic nature. To this end, we first sequenced the whole exomes of four PD patients carrying the LRRK2 G2019S mutation (L2-PD), seven idiopathic PD patients (ID-PD), three non-manifesting carriers of the LRRK2 G2019S mutation (L2-NMC), and three healthy individuals (controls). Variant calling in L2-NMC exomes identified 65,696 variants (**Fig. 3A**), consistent with previous estimates of individual genetic variation^32^. We then refined our search by selecting variants most likely to impact gene function, such as stop-gain or code-altering variants with a predicted deleteriousness score (CADD, see^33^) higher than 25, present in homozygosity or compound heterozygosity. This filter reduced the number of variants present in L2-NMC exomes to 421, of which 145 were exclusive to L2-NMC exomes (present in at least one L2-NMC exome and absent from all other groups; **Table S2**). Gene ontology analyses of the gene sets for which variants were present in L2-NMC exomes did not identify statistically significant enrichment in specific categories of molecular function, biological process, or cellular component (data not shown).

**Fig. 3.**
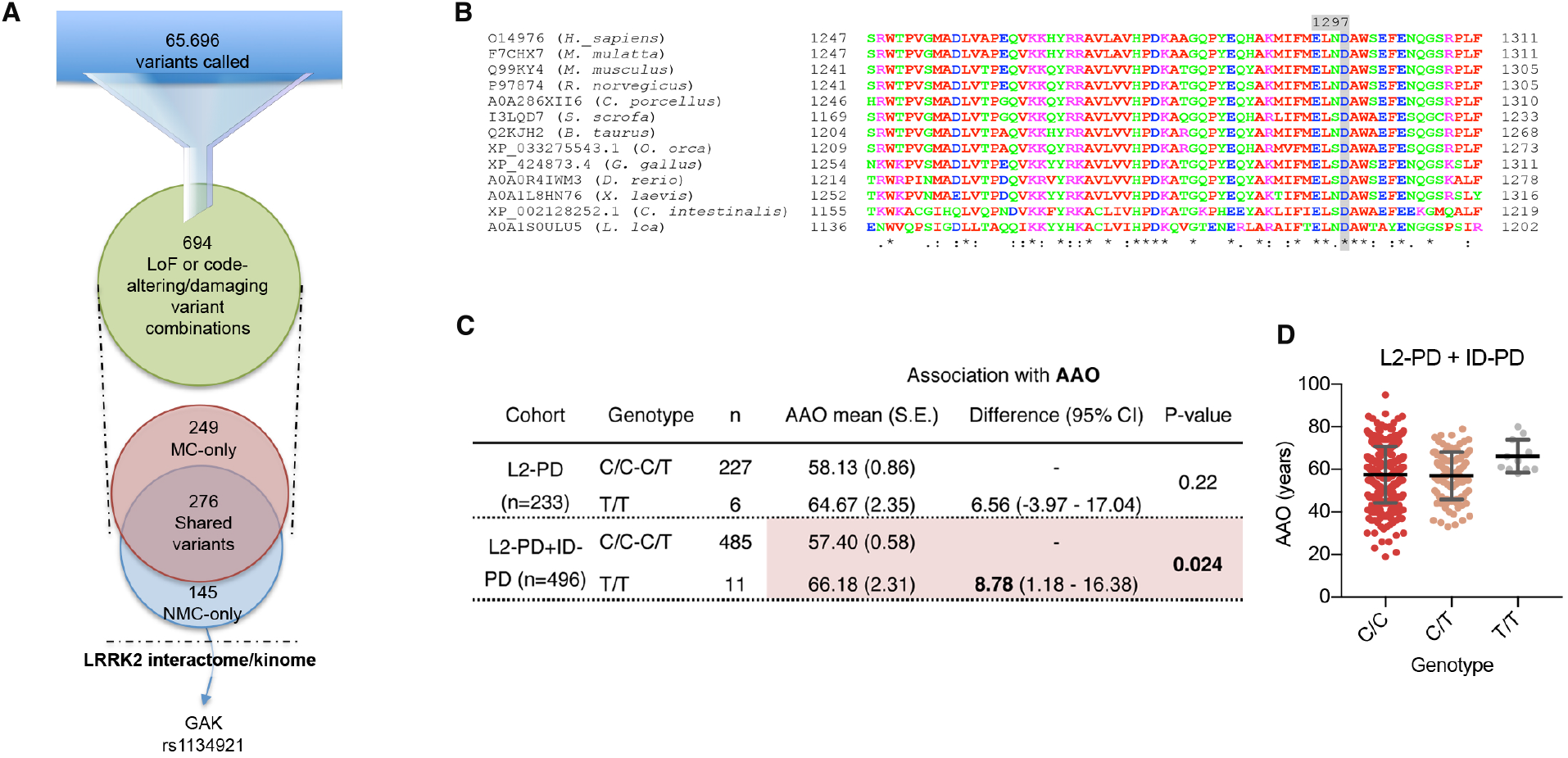
Exome interrogation reveals a L2-NMC-specifc variant that shows association with AAO in a local cohort. (**A**) Depiction of the strategy followed to filter the variants called in whole exomes (see main text for details). (**B**) Multiple sequence alignment of the J domain of GAK orthologs using Clustal Omega shows conservation of the human D 1297 residue across species. Proteins are identified by their UniProtKB ID or, when not available, by their NCBI Reference Sequence ID. (**C**) Association study of GAK rs11342921 C/T SNP and age-at-disease-onset (AAO) in a cohort of 233 L2-PD cases (top rows), or in combination with 263 cases of idiopathic PD (ID-PD, bottom rows). Only results from the recessive model yielded statistically significant results (p=0.024). (**D**) Dot plot showing the distribution of AAO of patients carrying each of the GAK rs1134921 genotypes. Asterisk denotes statistically significant differences (* p<0.05). [F(2,493) = 2.633; p=0.0729].

To begin unraveling the significance of genetic variants identified in L2-NMC exomes, we next applied a second filter for selecting variants located in genes previously associated with LRRK2 function^12,34–38^. Only one genetic variant could be recovered after applying this second filter; it corresponded to a single nucleotide variation (rs1134921, NC_000004.11:g.843508C>T; GRCh37.p13 chr 4) present in homozygosity in the exome of one of the L2-NMC subjects (codename SP_20). According to the Genome Aggregation Database (gnomAD v2.1.1^39^), this polymorphism is present in homozygosis in 2.00% of the overall population (2,812 out of 280,488 total alleles), with population frequencies ranging from the highest in the Latino population (5.56% or 979 out of 35,218 total alleles) to the lowest in the African population (0.13% or 16 out of 24,528 total alleles). The genetic variant rs1134921 results in a D1297N amino acid change in the J domain of the cyclin G-associated kinase (GAK). Multi-species protein sequence alignment indicates that the amino acid D1297 in GAK is highly conserved (Fig. 3B), thus suggesting functional importance. Nonetheless, the alternative variant is the reference one in the fruit fly, likely indicating that it is not a loss of function allele. To our knowledge, the GAK^D1297N^ isoform has not previously been characterized and neither has it been associated with biological or pathological processes.

### The GAK^D1297N^ variant association with age-at-disease-onset is heterogeneous across cohorts

To investigate the possible implication of the GAK^D1297N^ isoform in the manifestation of LRRK2 G2019S PD, we collected detailed clinical information from a total of 233 L2-PD patients followed at the Movement Disorders Unit of Hospital Clínic in Barcelona and genotyped them for the rs1134921 variant. Six of the 233 L2-PD patients (2.58%) carried the variant T polymorphism in homozygosis (T/T), whereas the remaining patients carried the reference allele or were heterozygous (C/C or C/T) for this SNP, consistent with estimates of 2.00% in the general population (gnomAD v2.1.1 database, see above). These genotype frequencies were not significantly different from those observed in a control cohort of 2,968 Spanish individuals (**Table S3**). A linear regression model was used to assess whether variation in age-at-disease-onset (AAO) of L2-PD patients could be explained by the different rs1134921 SNP genotypes. Although differences were not statistically significant, our analyses revealed a trend towards increased AAO in L2-PD patients homozygous for the T allele compared with those homozygous for the reference allele (C/C) or heterozygous (C/T) (**Fig. 3C**). To increase statistical power, we resorted to other cohorts to increase the numbers of patients homozygous for the T variant at the rs1134921. Since it has been reported that LRRK2 function can play important pathogenic roles even in ID-PD cases^40,41^, a second cohort of 263 ID-PD patients from the same clinic, with no mutations in LRRK2, was included in the analysis to increase the statistical power. When using this extended cohort, PD patients homozygous for the variant T at the rs1134921 SNP had a statistically significant delay of AAO of almost nine years compared with other genotypes at this SNP (**Fig. 3C-D**). We then sought to replicate our findings in three independent cohorts of LRRK2 G2019S carriers from diverse ancestries (French, North African and Ashkenazi Jewish). However, the remarkably low frequency of the T/T genotype in these cohorts (0.6% in the combined dataset), precluded conducting any association study in these populations.

These numbers give an idea of how difficult is to gather enough statistical power to conduct association studies with rare or quasi-rare genotypes; this is especially the case in genetic studies concerning age at disease onset given the subjective nature of this measure^42^.

### Controlling the genomic background confirms GAK^D1297N^ isoform protective effect in dopamine neurons

To circumvent the difficulty of replicating these genetic findings in independent cohorts, we resorted to CRISPR/Cas9-mediated genome editing to generate isogenic iPSC lines differing only in the GAK isoform in two unrelated genetic backgrounds (Fig. S4). Thanks to the isogenic genetic background, we can study the effect of the variant in isolation. Interestingly, the increased numbers of apoptotic DAns seen in L2-PD cultures were completely reverted simply by substituting the common GAK isoform by the GAK^D1297N^ isoform (**Fig. 4 A-B**). Conversely, replacing the GAK^D1297N^ isoform in L2-NMC DAns with the common GAK isoform rendered these cells highly susceptible to neurodegeneration, reaching levels of DAn apoptosis comparable to those of L2-PD cultures (**Fig. 4 A-B**). These results show that the relative protection from *in vitro* neurodegeneration observed in DAn cultures of the L2-NMC SP_20 background is caused by the expression of the GAK^D1297N^ isoform in this individual, and they suggest that carrying this low-frequency variant in GAK may be protective against PD onset and/or progression.

**Fig. 4.**
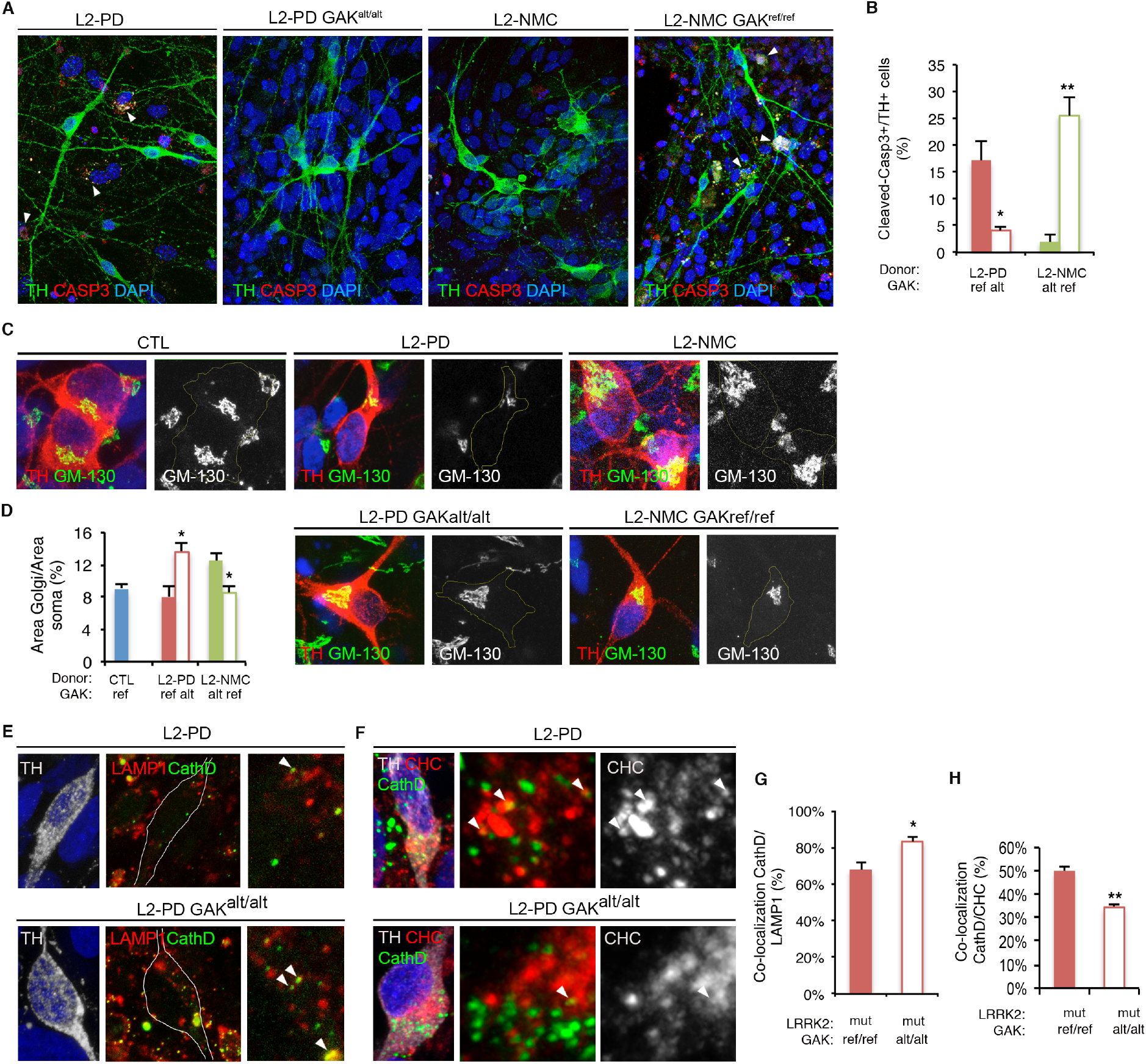
GAK^D1297N^ protects against neurodegeneration *in vitro*. (**A**) Immunofluorescence analyses of DAns differentiated from iPSCs of the indicated genotypes after 75 days of culture on top of mouse cortical astrocytes, stained for tyrosine hydroxylase (TH, green), the apoptotic marker cleaved caspase-3 (Casp3, red), and DAPI (blue). Arrows indicate TH+/Casp3+ cells. (**B**) Quantitative analysis of the percentage of apoptotic DAns at day 75. Data are averages ± s.e.m. of at least three independent experiments. L2-PD-GAK^ref/ref^, 169 DAns from SP_12; L2-PD-GAK^alt/alt^, 401 DAns from SP_12-GAK^alt/alt^; L2-NMC-GAK^ref/ref^, 292 DAns from SP_20-GAK^ref/ref^; L2-NMC-GAK^alt/alt^, 96 DAns from SP_20. Asterisks denote statistically significant differences (* p<0.05; ** p<0.01). [F(3,8) = 19.08; p=0.0005]. (**C**) Immunofluorescence analysis of DAns differentiated from isogenic iPSC lines (originally derived from L2-PD SP_12) differing in the presence of the LRRK2^G2019S^ mutation or the GAK^D1297N^ variant, stained for TH (red) and cis-Golgi marker GM130 (green). Nuclei were counterstained with DAPI (blue). The green channel is shown to the right as grayscale with the DAn soma outlined by yellow lines to help visualization. (**D**) Quantitative analysis of the Golgi area related to the area of the DAn soma of the indicated genotypes. Data are averages ± s.e.m. of three independent experiments. Control (CTL; SP_11), 27 DAns; L2-PD (SP_12), 31 DAns; L2-PD GAK^alt/alt^, (SP_12-GAK^alt/alt^), 33 DAns. Asterisks denote statistically significant differences (*p<0.001). [F(2,6) = 6.729; p=0.0293]. (**E**) Immunofluorescence analysis of DAns of the indicated genotypes (parental iPSC line: L2-PD SP_12), stained for TH (gray), cathepsin D (CathD, green), and the lysosomal marker LAMP1 (red). Nuclei were counterstained with DAPI (blue). The boxed areas are magnified to the right and arrowheads point to cathepsin D-positive puncta co-localizing with the lysosomal membrane marker LAMP1. (**F**) Immunofluorescence analysis of DAns of the indicated genotypes (parental iPSC line: L2-PD SP_12), stained for TH (gray), cathepsin D (CathD, green), and the marker from recently evaginated vesicles clathrin-heavy chain (CHC, red). Nuclei were counterstained with DAPI (blue). Arrowheads on the gray scale image to the right point to CHC staining co-localizing with cathepsin D-positive puncta. (**G**) Quantitative analysis of the co-localization of cathepsin D-positive puncta with LAMP1 puncta shown in **E**. Data are averages ± s.e.m. of a minimum of three independent experiments. L2-PD, 21 DAns; L2-PD-GAK^alt/alt^, 21 DAns. Asterisk denotes statistically significant differences (*p<0.05). (**H**) Quantitative analysis of the co-localization of cathepsin D-positive puncta with CHC as shown in **F**. Data are averages ± s.e.m. of a minimum of three independent experiments. L2_PD, 17 DAns; L2-PD GAK^alt/alt^, 21 DAns. Asterisk denotes statistically significant differences (*p<0.005).

### The GAK^D1297N^ isoform improves endocytic trafficking of relevant cargoes

GAK is a multidomain protein member of the DnaJ homolog C (DNAJC) family^43,44^, primarily localized to the trans-Golgi network of most types of cells, where it plays essential roles in clathrin-mediated transport^43,45–47^. Overexpression of the common isoform of GAK in primary neurons or cell lines induces dramatic changes in Golgi morphology, which becomes clustered or displays minimal staining^36^. Conversely, depletion of GAK results in Golgi cisternae being fragmented, distended, or elongated in hepatocytes^46^ and HeLa cells^43^. Given the reported role of GAK in maintaining Golgi morphology, we used the Golgi-stack marker GM130 to stain dermal fibroblasts from a healthy donor and a L2-PD patient, both carrying the common GAK isoform, and from the L2-NMC individual carrying the GAK^D1297N^ isoform (**Fig. S5A**). While the Golgi apparatus was well developed in fibroblasts from all three sources, its size was significantly larger in L2-NMC fibroblasts carrying the GAK^D1297N^ isoform (**Fig. S5B**). These differences were much more evident in iPSC-derived DAns from the same individuals; control DAns showed a well-developed Golgi in close proximity to the cell nucleus, which contrasted with the clustered GM130 staining farther away from the nucleus of L2-PD DAns, whereas the Golgi in L2-NMC DAns (expressing the GAK^D1297N^ isoform) was remarkably larger than that of any other group (**Fig. 4C**). Replacing the common isoform of GAK in L2-PD DAns for the GAK^D1297N^ isoform resulted in better developed and significantly larger Golgi apparatus, whereas reverting the GAK genetic variant of L2-NMC DAns back to the reference allele led to small and clustered Golgi (**Fig. 4C-D**). These results confirm previous studies linking GAK function with Golgi morphology and structure ^36,46,48^ and show that the GAK^D1297N^ isoform is associated with larger and apparently better developed Golgi apparatus in DAns.

To investigate whether the association of the larger Golgi with the GAK^D1297N^ isoform had functional consequences, we evaluated clathrin-mediated vesicle transport from the *trans*-Golgi network by analyzing the trafficking of cathepsin D to lysosomes. For this step, we again generated DAns from L2-PD-GAK^ref/ref^ and from L2-PD-GAK^alt/alt^ iPSC lines and quantified the co-localization of cathepsin D with the lysosomal marker LAMP1. While the numbers of cathepsin D-positive puncta were similar in either type of cell, DAns expressing the GAK^D1297N^ isoform showed an increased co-localization of cathepsin D-positive puncta with LAMP1-positive vesicles, compared with L2-PD-GAK^ref/ref^ DAns expressing the common GAK isoform (**Fig. 4E, G**). Moreover, we noted a reduced co-localization of cathepsin D-positive puncta with clathrin-coated vesicles in L2-PD-GAK^alt/alt^ DAns compared with L2-PD-GAK^ref/ref^ DAns (Fig. 4F, H). Taken together, these results show that the GAK^D1297N^ isoform is associated with improved endocytic trafficking in DAns and suggest that the mechanism by which the GAK^D1297N^ isoform enhances trafficking of cathepsin D towards the lysosome may involve an acceleration in the intermediate/clathrin dissociation step.

## Discussion

Unraveling the genetic bases of complex diseases such as PD presents a phenomenal challenge. Even for the approximately 5% of cases considered to be monogenic familial cases, in which a driver mutation confers extremely high risk of suffering the disease, clinical features vary widely among patients and knowledge of the mutation has little bearing on individual prognosis or treatment choice^3^. The importance of genetic modifiers to explain variability in clinical presentation is supported by numerous studies^49,50^, yet their identification is hampered by the small sample size available for association analyses and the relative low weight of individual modifiers in the overall genetic risk^51^. Our study demonstrates that the integration of precisely selected patient cohorts—defined by their clinical and genetic profiles ^52^ —with advanced cellular modeling and genome editing technologies can uncover protective genetic variants, even when working with limited sample sizes.

Induced pluripotent stem cell (iPSC)-based models capture the full genetic complexity of human individuals and allow disease-relevant phenotypes to be observed in a controlled environment, minimizing the influence of external factors. When paired with precise genome editing, this strategy allows for functional testing of causality rather than mere association. In our stud we found evidence of genetic protection against PD in three individuals, and interrogation of their exome identified the specific genetic modifier responsible for the protection in one of them. We believe that individual SP_20 may have been the easiest to dissect with our experimental design because this subject showed the strongest genetic protection (Fig. 2B), which turned out to depend mostly on a single coding variant. Future investigation will be necessary to pinpoint the exact nature of the genetic protection in the remaining two individuals, which would have been missed in this first study if they consisted on non-coding variants^53^ and/or affected genes not previously associated with LRRK2 function.

Our cumulative results identified the GAK^D1297N^ isoform, homozygous in ∼2% of the population, as conferring protection against PD. This finding is consistent with previous GWAS that found non-coding polymorphisms in the *GAK* region (rs11248060>T and rs1564882>T) associated with reduced *GAK* expression in brain and increased risk for familial PD^54,55^. Mechanistic insights into the involvement of the common GAK isoform in PD pathogenesis came from findings of genetic interaction with alpha-synuclein^55^ and its direct binding to LRRK2 to promote Golgi clearance^36^. We show that these mechanisms are related and also underpin the protective effects of the

GAK^D1297N^ isoform found here which, compared with the common GAK isoform, enhanced trafficking of cathepsin D towards the lysosome. Considering that the variation in GAK identified here affects the J domain, responsible for stimulating the ATPase activity of Hsc70 during clathrin disassembly^56^, we hypothesize that the D1297N substitution improves overall vesicle trafficking by speeding up vesicle uncoating. In this respect, *in silico* simulations of the interaction between the J domain of auxilin and the nucleotide-binding domain of Hsc70 revealed that the equivalent D residue in bovine auxilin (D896) was involved in such an interaction^57^. This protective mechanism is in line with previous reports pointing at disrupted vesicle trafficking and impaired endocytosis as major pathological consequences of LRRK2 mutations ^35,58,59^.

Clathrin-mediated transport is among the cellular pathways most recurrently found to be altered in genetic studies of PD^59,60^. Particularly relevant to our findings, some genes in this pathway have been previously associated with PD AAO in GWAS of familial and LRRK2 PD^12,61^, including intronic variation in the region containing *AAK1*^61^. *AAK1* encodes AP2-associated kinase 1, which contains a serine/threonine-kinase domain related to that of GAK, and with which it shares phosphorylation substrates^62^. However, no association with the rs1134921 SNP identified in our studies was detected in previous PD AAO GWAS^63,64^, nor in the more recent LRRK2 PD AAO GWAS^65^, where, in contrast, a previously reported SNP in the *GAK* gene locus did reach nominal significance. We believe that this could be due to most recent GWAS having investigated association mainly under additive models, whereas our data show that rs1134921 C>T only confers protection when present in homozygosis.

An additional consideration is population heterogeneity. Previous reports have highlighted significant variability in modifier effects between ethnic groups—for example, DNM3 rs2421947 in Arab-Berber carriers of LRRK2 G2019S^1 2^. In contrast, our cohort was drawn largely from a single clinical center, using consistent diagnostic and inclusion criteria. This relative homogeneity may have improved signal detection. Still, the protective genotype (homozygous rs1134921 TT) was rare among LRRK2 carriers (<1%), which limited our ability to replicate findings in additional cohorts. This again reflects a broader challenge of studying low-frequency variants in rare subgroups, and emphasizes the need for international collaboration and meta-analytic approaches.

Moreover, our recruitment strategy focused on individuals with genetic risk factors and comprehensive prodromal assessment—criteria typically available only in large, well-characterized prospective cohorts such as the Parkinson’s Progression Markers Initiative (PPMI)68. Despite these efforts, we identified a protective coding variant in only one of three such individuals. While this underscores the potential of our approach, it also highlights its limitations. Expanding this pipeline will require broader inclusion of high-risk but asymptomatic individuals, integration of multi-omic datasets (e.g., genome, transcriptome, chromatin accessibility), and application of machine learning to identify subtle, polygenic modifiers6?,^7°^.

In summary, our findings offer mechanistic insight into how variation in GAK modulates cellular vulnerability in PD, reinforcing the importance of vesicle trafficking and lysosomal function in disease pathogenesis. More broadly, this work provides proof-of-concept that protective genetic factors—often overlooked—can be uncovered through thoughtful cohort selection and functional modeling, opening new avenues for prognosis, patient stratification, and ultimately, therapeutic intervention.

## Data Availability

Ethical and legal approval to carry out the ERC Starting Grant project (n. 311736), which included the derivation of three independent iPSC lines from three non-manifesting carriers recruited at the Hospital Clinic of Barcelona, was obtained from the Departament de Salut of the Generalitat de Catalunya on April 17, 2013.

## Acknowledgments

The authors are indebted to the PD patients, non-manifesting carriers, and healthy individuals who agreed to participate in this study. The authors thank Xavier Muñoz and Marta Pineda, from the Hereditary Cancer group at IIDBELL, for helpful advice on analysis of polymorphisms, and all members of the P-CMR[C] for productive discussions. We are grateful to the Advanced Fluorescence Microscopy Unit of the Institute of Biomedicine of the University of Barcelona (especially to Elena Rebollo Arredondo).

## Author contributions

Conceptualization, E.T., A.R., and A.C.; Methodology, C.C., M.C., V.M., C.M., T.C., F.H.G., A.R., and A.C.; Investigation, C.C., I.F.-C., Y.R.-P-, A.F., S.L., and C.M.; Formal analysis, N.S., R.F.-S., S.B.C., T.C. and M.E.; Resources, A.M., J.M.C., A.G., M.J.M, and E.T.; Supervision, J.M.C., V.M., E.B., T.C., A.R., and A.C.; Writing - Original Draft, C.C.; Writing – Review & Editing, A.R. and A.C.; Funding acquisition, A.R. and A.C.; Project administration, A.C.;

## Data and materials availability

All raw data are available in the manuscript.

## Supplementary Materials

Materials and Methods

Figures S1-S5

Tables S1-S3

References (*1-13*)

