## Supplemental Information for "Low-frequency genetic variants in GAK enhance Golgi function and protect against Parkinson’s disease"

**This PDF file includes:**

Materials and Methods
Figs. S1 to S5
Table S1 and S3
References

### Materials and Methods

Recruitment of non-manifesting carriers of the LRRK2 G2019S mutation and study of the prodromal features

Studies were approved by the authors' Institutional Review Board and conducted under the Declaration of Helsinki. Patients were encoded to protect their confidentiality, and written informed consent was obtained. The generation of human iPSCs was done following a protocol approved by the Spanish competent authorities (Commission on Guarantees concerning the Donation and Use of Human Tissues and Cells of the Carlos III Health Institute). Non-manifesting carriers were recruited from families having members affected by LRRK2-associated PD who attended the Movement Disorder Unit at the Hospital Clinic of Barcelona (Barcelona, Spain). They were selected on the basis of having an advanced age and little or no prodromal signs at the moment of enrollment.

Generation of iPSCs

Using the CytoTune iPSC Sendai reprogramming protocol, we converted fibroblasts into transgene-free iPSCs. Briefly, explant cultures were obtained from skin punch biopsies. Primary cultures of fibroblast were expanded and 50,000 to 100,000 cells were transduced with the Sendai vectors. Medium was then shifted to human embryonic stem cell (hESC) medium, consisting of KO-DMEM (Invitrogen) supplemented with 20% KO-Serum Replacement (Invitrogen), 2 mM Glutamax (Invitrogen), 50 µM 2-mercaptoethanol (Invitrogen), non-essential amino acids (Lonza) and 10 ng/ml bFGF (Peprotech). Cultures were maintained at 37°C, 5% CO2, with media changes every other day. Colonies were picked based on morphology 20-30 days after the initial infection and plated onto fresh feeders. Lines of patient-specific iPSCs were maintained by mechanical dissociation of colonies and splitting 1:3 onto feeder cells in hESC medium or by dissociation with EDTA and passaging onto Matrigel-coated plates with hESC medium pre-conditioned by mouse embryonic fibroblasts (chESC medium).

Characterization of iPSCs

The expression of Sendai vector transgenes and endogenous pluripotency-associated transcription factors was assessed by quantitative Polymerase Chain Reaction (after reverse transcription) (RT-qPCR). *In vitro* differentiation towards endoderm, mesoderm and neuroectoderm was carried out essentially as previously described[1].

Generation of TALEN monomers, CRISPR/Cas9 plasmids, recombination reporters and donor templates for HDR

TALE-based DNA binding domains (DBDs) were engineered as described elsewhere[2] at the Institute for Transfusion Medicine and Gene Therapy (University of Freiburg). The DBD includes 17,5 repeat units each containing a repeat variable di-residue (RVD) that defines its nucleotide specificity. Units containing the NI, NG, HD and NN RVDs were used to recognize A, T, C and G nucleotides respectively. Repeat units were arranged in tandem to recognize a user-defined 19-bp genomic target sequence and each DBD was fused to the wild-type FokI nuclease domain resulting in TALE-nuclease (TALEN) monomers with different specificity. Each monomer was inserted in a plasmid under the control of a modified CMV promoter[3]. CRISPR/Cas9 plasmid pSpCas9(BB)-2A-GFP (PX458)[4] was a kind gift from Feng Zhang (Addgene plasmid #48138). Original pCbh promoter was exchanged for the full-length pCAGGS promoter to achieve higher expression levels in hiPSCs. Custom guide RNAs were cloned into the BbsI sites as annealed oligos. Donor templates for HDR were generated using standard molecular cloning procedures. Briefly, for the LRRK2 donor template, homology arms were amplified from genomic DNA from either wild-type or LRRK2 G2019S mutant hiPSC lines and inserted into the KpnI-XhoI (5'HA) and SpeI-NotI (3'HA) sites of pBS-SK(-). pRex1-NeoR-SV40pA cassette was amplified from aMHC-eGFP-Rex-Neo[5] (kind gift from Mark Mercola; Addgene plasmid #21229) with primers containing LoxP sites in the proper orientation for excision and inserted into the SalI-BamHI sites. Recombination reporter pRR-Puro was a kind gift from Marc Bühler (Addgene plasmid #65853)[6]. A 37 bp-long sequence comprising guide RNA target sequences were cloned in between SacI and AatII sites as annealed oligos.

Gene edition in iPSCs

To correct the LRRK2 G2019S mutation, mutant iPSCs were gene edited using TALENs. iPSCs grown to confluence in 10cm plates were pre-treated for 2-4 hours with 10 μM Y-27632 (RI; Miltenyi-Biotech), disaggregated to small clumps using Accutase (eBiosciences), resuspended in ice-cold chESC medium supplemented with RI and containing 15 μg of each TALEN monomer-coding plasmids and 30 μg HDR donor template and placed in a electroporation cuvette. Cells were electroporated with a Gene Pulser Xcell electroporation system (BioRad) with the following settings: 250 V and 500 μF (time constant should be between 10 and 14 milliseconds). After being pulsed, the cell suspension was seeded in 10-cm plates coated with Matrigel containing RI-supplemented chESC medium. Seventy-two hours post-transfection, 50 μg/mL G-418 (Melford Laboratories Ltd.) treatment was initiated and maintained for two weeks until resistant colonies attained enough size to be screened. At that moment, half of each resistant colony was manually picked and site-specific integration was verified by means of PCR. Gene correction was assessed by Sanger sequencing. Colonies with the desired genotype were isolated, expanded and cryopreserved.

For inserting the mutation in heterozygosis, wild-type iPSCs were edited using CRISPR/Cas9. This choice was made based on the difficulty of controlling the zigosity of the edition using TALENs. CRISPR guide RNAs overlapping the selection cassette insertion site were observed to favor biallelic integrations (data not shown). The day before transfection, 800,000 iPSCs were seeded on Matrigel-coated 10-cm plates. The next day, cells were transfected using FuGENE HD (Promega) and a 2:1:1 mixture of the following plasmids: a CRISPR/Cas9 plasmid encoding for a guide RNA whose spacer sequence overlapped selection cassette insertion site and two donor plasmids bearing either the wild-type or the G2019S alleles. Subsequent steps were carried out as described with TALENs.

For the excision of the selection cassette, edited iPSCs were transfected with CRE recombinase-expressing plasmid (kind gift from Michel Sadelain; Addgene plasmid #27546)[7]. Forty-eight hours post-transfection, cells were singularized and seeded at clonal density on a feeder layer of irradiated human fibroblasts. When colonies attained a certain size they were picked and subcultured in independent Matrigel-coated wells. Cells were sampled and checked for cassette excision by PCR and Sanger sequencing. Clones in which the cassette was excised were expanded, cryopreserved and karyotyped.

For the generation of GAK rs1134921 isogenic controls, selected iPSC lines were seeded at a density of 300,000 cells per well of a six-well plate. The next day, they were co-transfected using Lipofectamine Stem (ThermoFischer) and 1:1:2 mixture of the following plasmids: a CRISPR/Cas9 plasmid encoding for an allele-specific guide RNA, a pRR-Puro plasmid containing the corresponding guide RNA spacer sequence and a single-stranded oligodeoxynucleotide as donor template for HDR (ssODN) (SIGMA). Moreover, the ssODN contained two further nucleotide changes for facilitating genetic screening and for avoiding re-cleavage of edited alleles. Thirty-six hours after transfection, 1 μg/mL puromycin was added to the medium and maintained another 36 hours. Those colonies that resisted puromycin treatment were isolated and molecularly characterized by restriction site length polymorphisms and Sanger sequencing. Clones presenting bi-allelic recombination and the desired genotype were expanded, cryopreserved and karyotyped.

iPSC differentiation to DA neurons

For DAn differentiation, iPSCs were transduced with LV.NES.LMX1A.GFP and processed as previously described[8]. For DAn yield analysis cells were co-cultured with PA6 for three weeks in N2B27 medium. For short-term SNCA analysis, DAns generated on the top of PA6 for three weeks were trypsinized and cultured for three days on Matrigel-coated dishes. For long-term culture, neural progenitor cells were seeded onto mouse primary cortical astrocytes, prepared as described elsewhere[9], and maintained in N2B27 medium. After nine weeks, cells were fixed and processed for immunofluorescence analysis.

Immunofluorescence

Cells were fixed with 4% paraformaldehyde in PBS at room temperature (RT) for 20 min and permeabilized for 15 min in 0.3% Triton in TBS. Cells were then blocked in Triton-X100 with 3% donkey serum for two hours. The following antibodies were used: goat anti-Nanog (R&D Systems; AF1997; 1:50), mouse IgM anti-Tra-1-81 (Merck-Millipore; MAB4381; 1:200), mouse anti-OCT4 (Santa Cruz; sc-5279; 1:30), rat IgM anti-SSEA-3 (Developmental Studies Hybridoma Bank (DSHB); MC-631; 1:10), mouse-SOX2 (R&D Systems; MB2018; 1:50), mouse anti-SSEA-4 (Developmental Studies Hybridoma Bank (DSHB); MC-813-70; 1:100), mouse anti-TUJ1 (Biolegend; 801202; 1:500), rabbit anti-GFAP (Dako; Z0334; 1:1000), rabbit anti-AFP (Dako; A0008; 1:400), goat anti-FOXA2 (R&D Systems; AF2400; 1:50), mouse anti-SMA(Sigma; A5228; 1:400), rabbit anti-GATA4 (Santa Cruz; sc-9053; 1:50), rabbit anti-TH (Santa Cruz; sc14007; 1:500), sheep anti-TH (Pel-Freez P60101-0 1:500), rabbit anti-cleaved caspase-3 (Cell Signaling; 9664; 1:400), mouse anti SNCA (BD transduction laboratories; 610787; 1:500), rabbit anti-GM130 (BD transduction laboratories; 610822; 1:100), mouse IgM anti-Cathepsin D (Santa Cruz; sc-377299; 1:250), mouse anti-CHC (Santa Cruz; sc-271178; 1:50) and mouse anti-LAMP1 (Developmental Studies Hybridoma Bank (DSHB); H4A3; 1:100). Secondary antibodies used were all the Alexa Fluor Series from Invitrogen (all 1:200). Images were taken using Leica SP5 confocal microscope. To visualize nuclei, slides were stained with 0.5 μg/ml DAPI (4′,6-diamidino-2-phenylindole) and then mounted with PVA/DABCO.

Sholl analysis

Neurite morphology study was performed at the indicated time-points on iPSC-derived DAns differentiated on top of cortical mouse astrocytes fixed and immunostained for TH. We randomly selected fields from differentiated cultures and assessed neurite morphology. Z-stacks were used to unambiguously trace the neurites from the neuron being analyzed. Neurites were traced using the Simple Neurite Tracer plugin on FiJi and the neuronal complexity was measured by counting the number of neurite intersections with concentric circles radiating from the cell body with the Sholl Analysis plugin. The number of branch points and terminals was manually counted after the analysis.

Exome sequencing

A total of three healthy controls, seven sporadic PD patients, four LRRK2 G2019S PD patients described elsewhere[9] plus the three non-manifesting carriers of the LRRK2 G2019S described here (Table S1) participated in the genetic study. Genomic DNA was harvested from low passage dermal fibroblast using QIAamp mini DNA kit (Qiagen). Whole exomes were captured with the SureSelect V5 kit (Agilent) and were sequenced on an Illumina 2000/2500 instrument at the Genomics Unit from the Center for Genomic Regulation (Barcelona, Spain). Raw sequencing data quality was assessed using FASTQC and no relevant concerns were observed. The paired-end reads (read size: 125bp) were mapped to the human reference genome GRCh37 using bwa (version 0.5.9), allowing up to five mismatched, inserted or deleted bases (indels). The alignments were refined using GATK (version 1.6) by realigning reads around indels, removing PCR duplicates and performing base quality score recalibration. After the entire process, the actual mean base coverage of the samples was 34.09x. Variant calling was performed using the algorithm HaplotypeCaller from GATK and obtained variants were annotated using ANNOVAR[10].

Multiple protein alignment

Was done using Clustal Omega through the EBI Web services publicly available at www.ebi.ac.uk[11].

GO Enrichment analysis

Analysis was done using the PANTHER[12] Overrepresentation Test (Released 20200728), with GO Ontology database (Released 2020-08-10) and Homo sapiens (all genes in database) as reference list, with Fisher's Exact test with False Discovery Rate correction. Annotation datasets included PANTHER GO-slim Molecular Function, PANTHER GO-slim Biological Process, and PANTHER GO-slim Cellular Component.

Genotyping of candidate protective variants

Genomic DNA was extracted from peripheral blood following standard procedures. Genotyping was performed using a custom TaqMan assays for rs1134921 on a StepOnePlus Real-time PCR System (Applied Biosystems, Foster City, CA). Statistical analysis was performed using the SNPstats software (Sole et al. 2006). Linear regression models were used to assess the AAO variation explained by the different rs1134921 SNP genotypes under different possible inheritance models. Akaike's information criterion (AIC) and Bayesian information criterion (BIC) were calculated to define the data that best fitted the model. If binary, the application assumes an unmatched case–control design and unconditional logistic regression models are used.

Measurement of Golgi apparatus area

Dopaminergic neurons cultured for nine weeks on the top of cortical murine astrocytes were fixed and immunostained for the dopaminergic and the cis-Golgi markers TH and GM130. Dopaminergic neurons were imaged with a 63x objective and maximum projections were obtained. The area of the Golgi apparatus was obtained using FIJI and an in-house developed plug-in. The results were expressed as the fraction of the neuronal soma occupied by the Golgi apparatus.

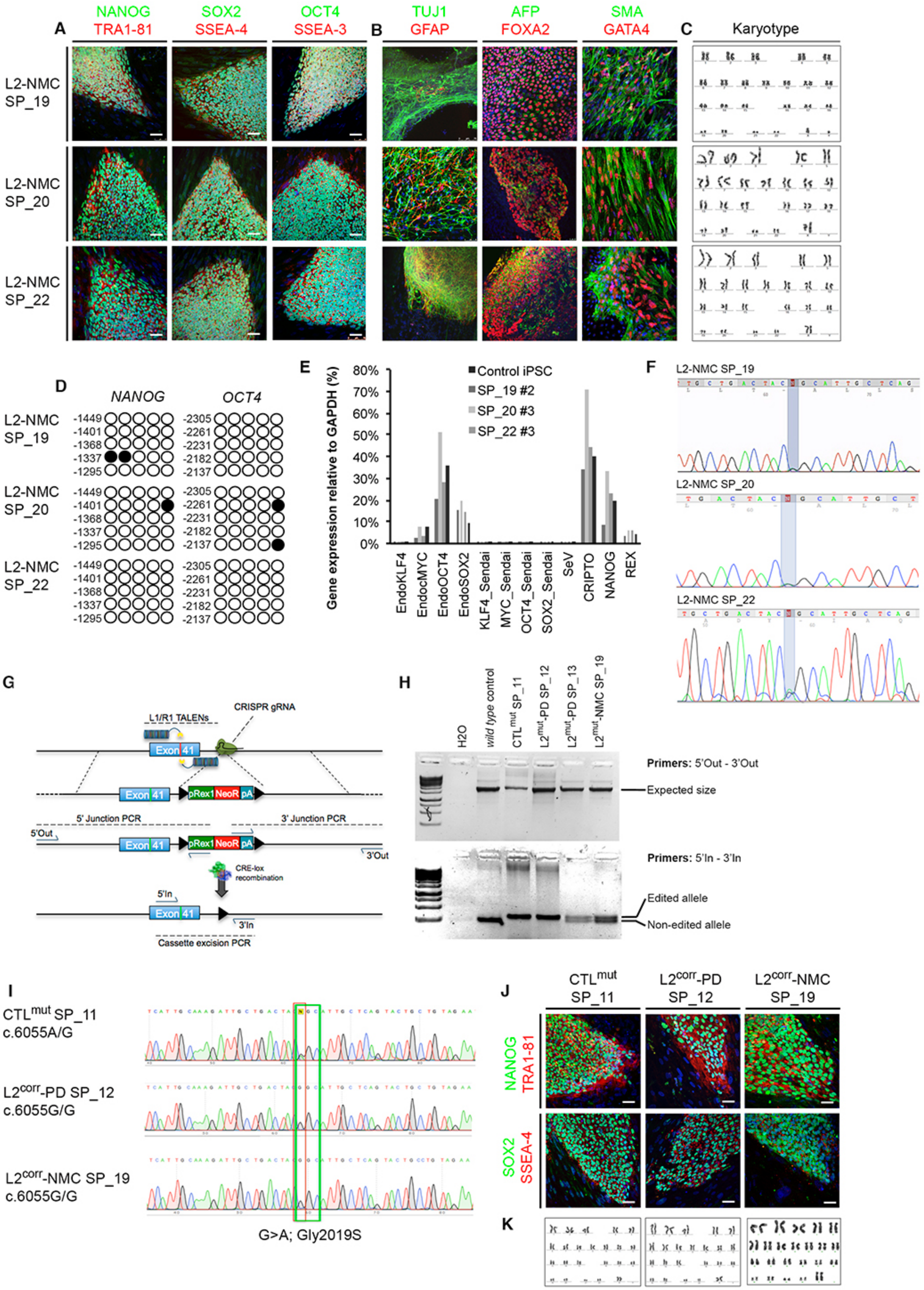
A
NANOG
TRA1-81
SOX2
SSEA-4
OCT4
SSEA-3
B
TUJ1
GFAP
AFP
FOXA2
SMA
GATA4
C
Karyotype
L2-NMC SP_19
L2-NMC SP_20
L2-NMC SP_22
D
NANOG
OCT4
E
Control iPSC
SP_19 #2
SP_20 #3
SP_22 #3
Gene expression relative to GAPDH (%)
EndoKLF4
EndocMYC
EndoOCT4
EndoSOX2
KLF4_Sendai
MYC_Sendai
OCT4_Sendai
SOX2_Sendai
SeV
CRIPTO
NANOG
REX
F
G
L1/R1 TALENs
CRISPR gRNA
Exon 41
pRex1 NeoR pA
5' Junction PCR
3' Junction PCR
5'Out
3'Out
CRE-lox recombination
5'In
3'In
Cassette excision PCR
H
H2O
wild type control
Primers: 5'Out - 3'Out
Expected size
Primers: 5'In - 3'In
Edited allele
Non-edited allele
I
CTLmut SP_11 c.6055A/G
L2corr-PD SP_12 c.6055G/G
L2corr-NMC SP_19 c.6055G/G
G>A; Gly2019S
J
CTLmut SP_11
L2corr-PD SP_12
L2corr-NMC SP_19
K

**Fig. S1. Generation of iPSC lines from L2-NMC as well as of isogenic controls from both carriers and non-carriers.** (**A**) Immunofluorescence analysis of representative colonies of L2-NMC SP_19, SP_20 and SP_22 stained positive for the pluripotency-associated markers NANOG, OCT4 and SOX2 (green); TRA-1-81, SSEA3 and SSEA4 (red). (**B**) Immunofluorescence analyses of L2-NMC iPSC lines differentiated *in vitro* show the potential to generate cell derivatives of all three primary germ cell layers, including ectoderm (stained for TUJ1, green and GFAP, red), endoderm (stained for α-fetoprotein. green. and FOXA2. red) and mesoderm (stained for smooth muscle actin. SMA. red). (**C**) Normal karyotype from selected L2-NMC iPSC clones. (**D**) Bisulphite genomic sequencing of the NANOG and OCT4 promoters showing demethylation in L2-NMC iPSC lines. (**E**) RT-qPCR analyses of the expression levels of Sendai Virus-derived reprogramming factors and endogenous expression levels (Endo) of the indicated genes in L2-NMC iPSC lines and a previously validated iPSC line. (**F**) Sanger sequencing of LRRK2 exon 41 revealed the presence of the G2019S in heterozygosis. (**G**) Scheme describing the recombination steps given during the edition process. Blue arrows represent the primers used for the PCR screening procedure. Black triangles represent LoxP sites surrounding the selection cassette. Green and red bar in Exon 41 represents the wild-type and the G2019S alleles, respectively. (**H**) Molecular analysis of the correctly edited clones. Upper gel demonstrates the preservation of locus integrity using primers outside the sequence covered by the homology arms. Lower gel shows successful pRex1-NeoR cassette excision and allelic dose of the gene edition (upper band corresponds to the edited allele carrying the remaining LoxP; the lower band corresponds to the non-edited allele). (**I**) Sanger sequencing of LRRK2 exon 41 confirming the successful edition of LRRK2 c.60055 site. (**J**) Immunofluorescence analysis of representative colonies of isoCTL SP_11, iso-L2-PD SP_12, iso-L2-PD SP_13 and iso-L2-NMC SP_19 stained positive for the pluripotency-associated markers NANOG and SOX2 (green) and TRA-1-81 and SSEA4 (red). (**H**) Normal karyotype from selected gene-edited iPSC clones.

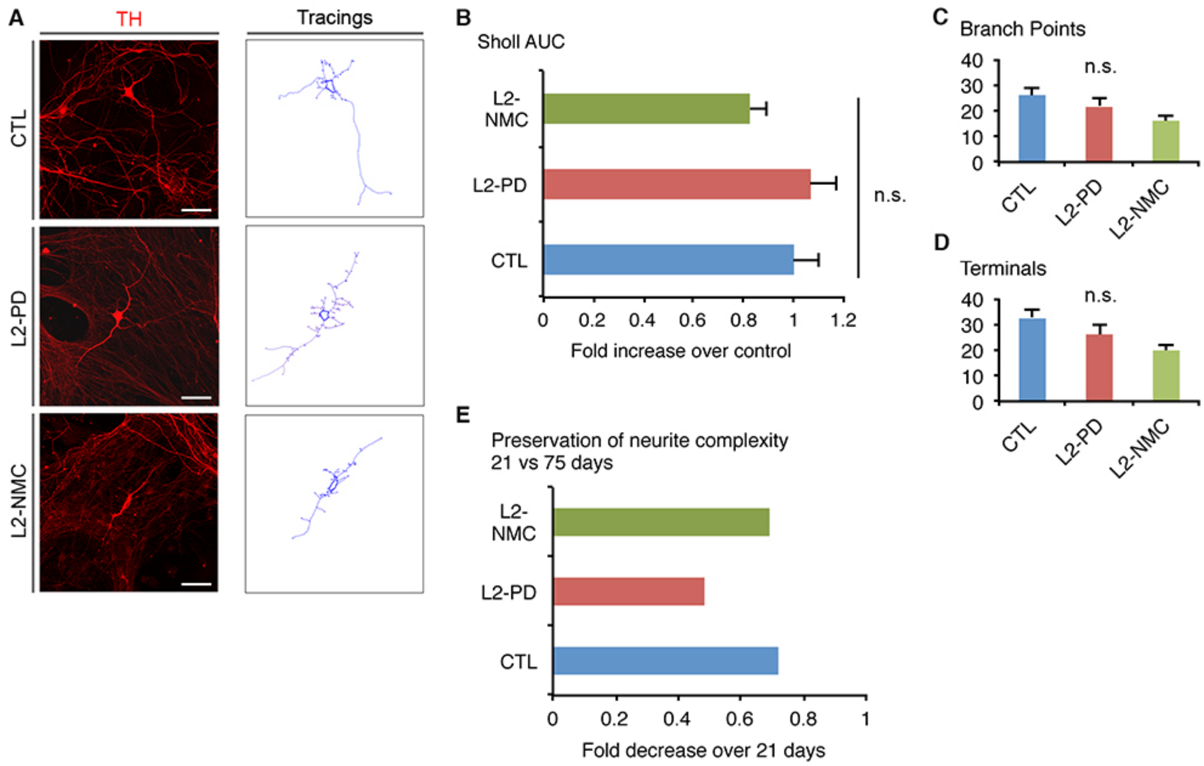


**Fig. S2. Morphological features of DAns after 21 days of culture.** (**A**) Immunofluorescence analysis of DAns differentiated from iPSC lines of the indicated genotypes and cultured for 21 days on top of mouse cortical astrocytes, stained for TH (red). Right panels show neurite tracings of representative DAns for subsequent Sholl analysis. Scale bar, 50 µm. (**B-D**) Quantitative analysis of the area under the curve (AUC) of the Sholl analysis (**B**), number of branch points (**C**), and number of terminals (**D**) performed in DAns differentiated from iPSC lines of the indicated genotypes and cultured for 21 days on top of mouse cortical astrocytes. Data are averages ± s.e.m. of at least two independent experiments: Control (CTL), 20 DAns from SP_11; L2-PD, 19 DAns from SP_12; L2-NMC, 34 DAns from 2 iPSC lines (SP_19 and SP_20). ANOVA for Sholl AUC: [F(2,70) = 2.416; p=0.0967]; for number of branches: [F(2,70) = 4.083; p=0.0210]; for number of terminals: [F(2,70) = 5.106; p=0.0085]. (**E**) Ratio of AUC values calculated at 21 days of culture (**Fig. S2B**) and at 75 days of culture (**Fig. 2E**) for DAns of the indicated genotypes.

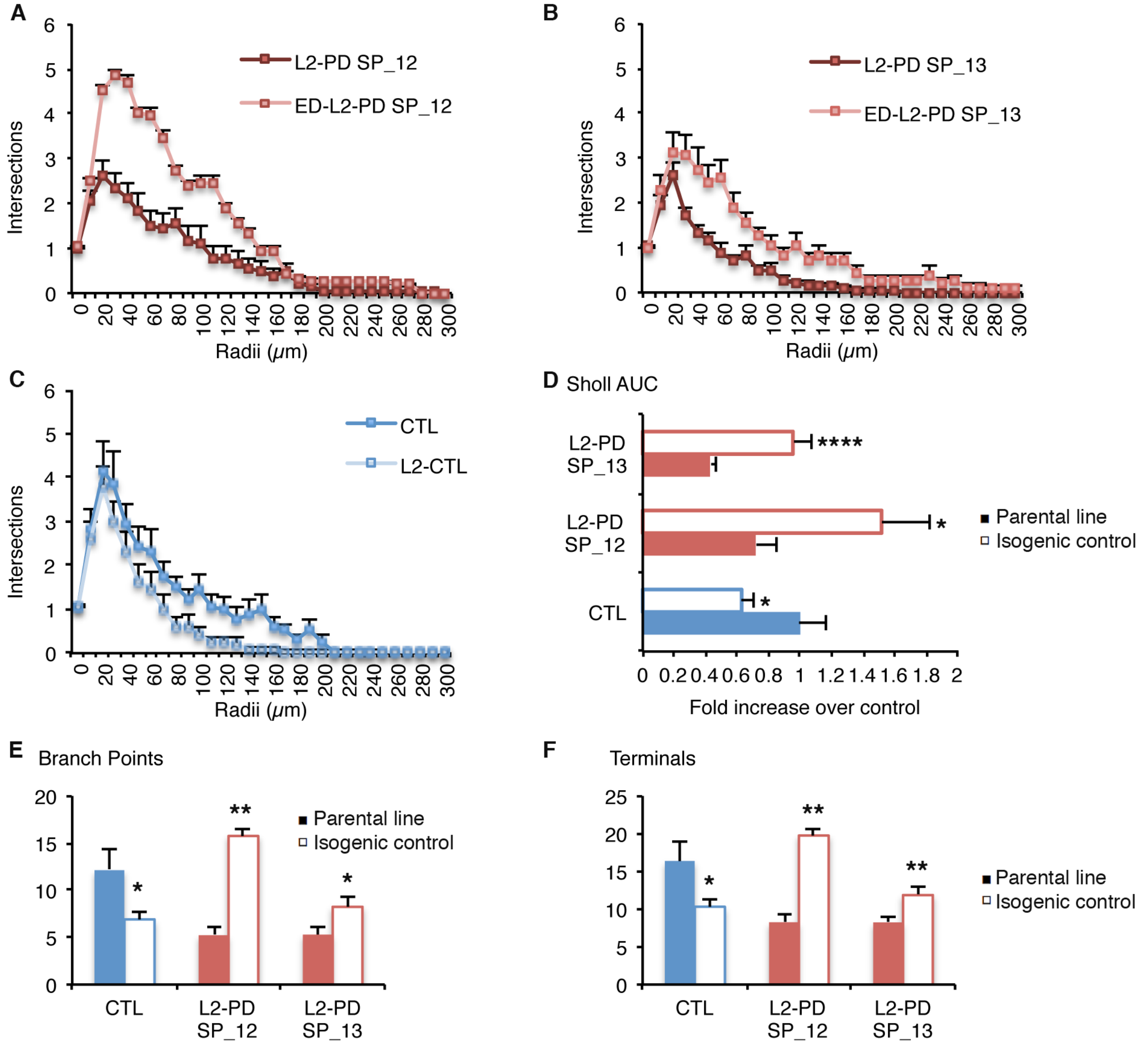


**Fig. S3. Morphological differences among isogenic lines after 75 days of culture.** (**A-C**) Sholl analysis of DAns differentiated from the parental iPSC line as well as from their LRRK2 G2019S genetically matched control from L2-PD SP_12 (**A**), L2-PD SP_13 (**B**) and Ctrl SP_11 (**C**) after 75 days on the top of cortical astrocytes. (**D-F**) Quantitative analysis of the area under the curve (AUC) of the Sholl analysis (**D**), number of branch points (**E**) and number of terminals (**F**) performed in DA neurons differentiated from three genetically matched iPSC lines and cultured for 75 days on the top of cortical astrocytes. Data are averages ± s.e.m. of at least two independent experiments. Control (CTL), 29 DAns from SP_11; $CTL^{mut}$ 29 DAns from $SP\_11^{mut}$; L2-PD SP_12, 30 DAns; L2-PD $SP\_12^{corr}$, 25 DAns; L2-PD SP_13, 33 DAns; L2-PD $SP\_13^{corr}$, 24 DAns. ANOVA for Sholl AUC: [$F(5,164) = 6.038$; $p<0.0001$]; for number of branches: [$F(5,163) = 6.618$; $p<0.0001$]; for number of terminals: [$F(5,163) = 6.422$; $p<0.0001$].

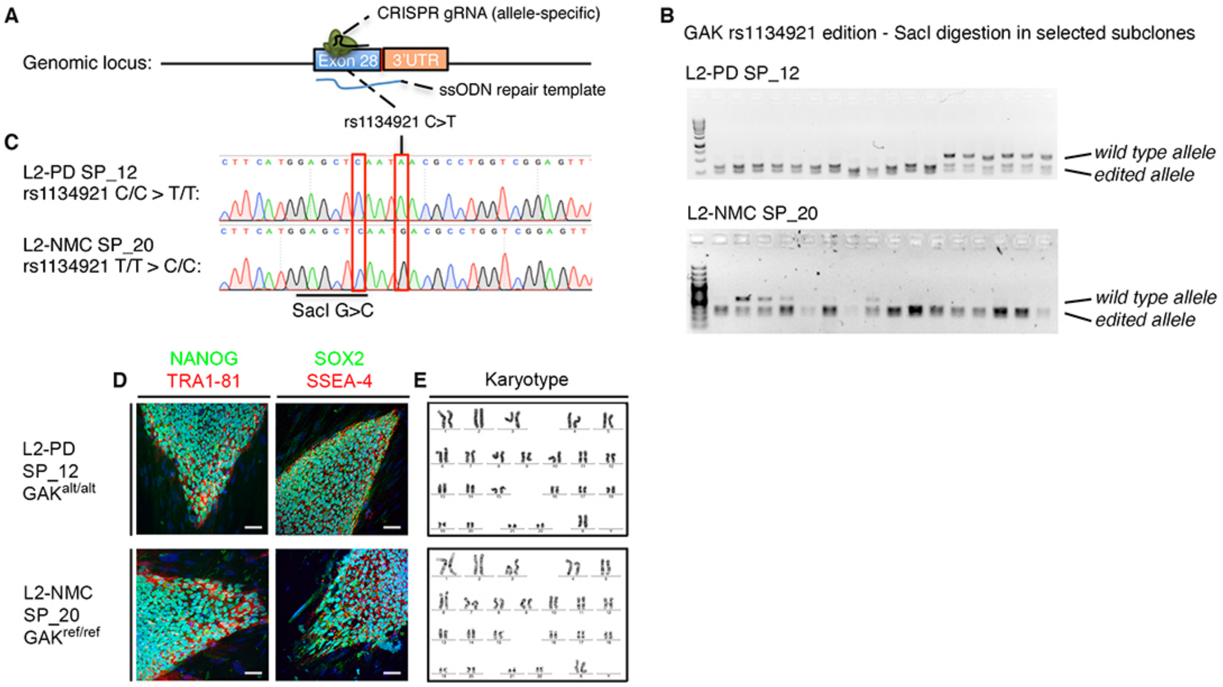


**Fig. S4. Generation of isogenic clones differing in the GAK rs1134921 variant. (A)** Scheme describing the editing of the rs1134921 genetic variant using CRISPR/Cas9 and a single-strand oligodeoxynucleotide (ssODN) as HDR template. ssODN harbors a silent base-pair change that introduces a SacI diagnostic site for subsequent molecular screening through RFLP. **(B)** Molecular analysis of the puromycin-resistant clones (see Methods section) through RFLP. Upper band is non-recombined allele; the lower ones correspond to recombined alleles cut with SacI restriction enzyme. **(C)** Sanger sequencing of GAK exon 28 confirming the successful edition of GAK rs1134921 site and the introduction of the SacI diagnostic site. **(D)** Immunofluorescence analysis of representative colonies of L2-PD SP_12$^{alt/alt}$ and L2-NMC SP_20$^{ref/ref}$ stained positive for the pluripotency-associated markers NANOG and SOX2 (green) and TRA-1-81 and SSEA4 (red). **(E)** Normal karyotype from selected gene-edited iPSC clones.

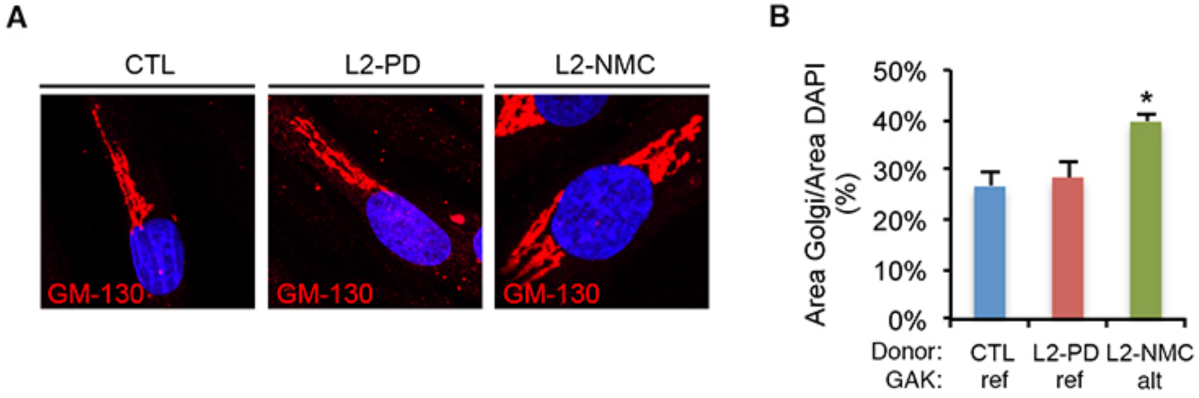


**Fig. S5. Golgi morphology in primary fibroblasts from LRRK2 manifesting and non-manifesting carriers.** (**A**) Immunofluorescence analysis of primary fibroblasts from CTL, L2-PD and L2-NMC lines, stained for the cis-Golgi marker GM130 (red). Nuclei were counterstained with DAPI (blue). (**B**) Quantitative analysis of the Golgi area related to the area of nuclei. Data are averages ± s.e.m. of two independent experiments: Control (CTL; SP_11), 10 cells; L2-PD (SP_12), 10 cells; L2-NMC (SP_20), 10 cells. Asterisks denote statistically significant differences (*$p<0.01$). [$F(2,26) = 10.17$; $p=0.0005$].

**Table S1. Summary of iPSC lines used in the current studies**

| Cell Line Code | Disease status | Gender | LRRK2 status | GAK status | Source |
|---|---|---|---|---|---|
| L2-NMC SP_19 | NMC | Male | G2019S/WT | ref/ref | Here |
| L2-NMC SP_19$^{corr}$ | " | " | WT/WT | " | Here |
| L2-NMC SP_20 | NMC | Female | G2019S/WT | alt/alt | Here |
| L2-NMC SP_20$^{ref/ref}$ | " | " | G2019S/WT | ref/ref | Here |
| L2-NMC SP_22 | NMC | Female | G2019S/WT | ref/ref | Here |
| CTL SP_11 | Control | Female | WT/WT | ref/ref | Ref.[9] |
| CTL SP_11$^{mut}$ | " | " | G2019S/WT | " | Here |
| CTL SP_17 | Control | Male | WT/WT | ref/ref | Ref.[9] |
| L2-PD SP_05 | PD | Male | G2019S/WT | ref/ref | Ref.[9] |
| L2-PD SP_12 | PD | Female | G2019S/WT | ref/ref | Ref.[9] |
| L2-PD SP_12$^{corr}$ | " | " | WT/WT | " | Here |
| L2-PD SP_12$^{alt/alt}$ | " | " | G2019S/WT | alt/alt | Here |
| L2-PD SP_13 | PD | Female | G2019S/WT | ref/ref | Ref.[9] |
| L2-PD SP_13$^{corr}$ | " | " | WT/WT | " | Ref.[13] |

Main characteristics of the iPSC lines used, including the disease status of the donors (NMC: non-manifesting carrier, PD: Parkinson's disease patient), gender of the donors, as well as the status of the *LRRK2* and *GAK* genes (WT: wild-type allele; ref: reference allele; alt: alternative allele), and source of the iPSC line, specifying those that were generated for this publication (Here).

**Caption for Table S2. Gene variants exclusive to NMC exomes**

Gene variants or variant combinations likely affecting the function of both copies of their corresponding gene and found exclusively in L2-NMC exomes and not in L2-PD, ID-PD, or control exomes.

Chr: Chromosome; Var(1/2) Type: Predicted functional consequence; Var(1/2) ID: Reference SNP code; Var(1/2) Position: Genomic positions according to the hg19/GRCh37 human assembly; Var(1/2) Ref./Alt. Allele: Sequence of the reference and alternative alleles, respectively; Var(1/2) CADD: Predicted deleteriousness score of the gene variant; Var(1/2) Genotype: Encoded as 0/1 for heterozygosity and 1/1 for homozygosity.

**Table S3. Association of GAK rs1134921 with PD risk under a recessive model**

| Cohort | Genotype | n | Frequency | OR (95% CI) | P-value (vs. control) |
|---|---|---|---|---|---|
| Control (n=2,968)* | C/C-C/T | 2,925 | 98.55% | - | - |
| | T/T | 43 | 1.45% | | |
| L2-PD (n=233) | C/C-C/T | 227 | 97.42% | 1.80 (0.76-4.27) | 0.18 |
| | T/T | 6 | 2.58% | | |
| L2-PD+ID-PD (n=496) | C/C-C/T | 485 | 97.8% | 1,54 (0.79-3.01) | 0.20 |
| | T/T | 11 | 2.2% | | |

* The genotype data from the control cohort was obtained from the Spanish National Bank of DNA using the Axiom Spain Biobank Array. OR: odds ratio; CI: confidence interval.
